# Self-health Management and Role of Nurses of Diabetic Patients: A systematic review during COVID-19 Pandemic

**DOI:** 10.1101/2025.05.12.25327369

**Authors:** Yan Bo

## Abstract

**Aims:** The study aims to take the COVID-19 pandemic as an example to provide a scientific reference for diabetic patients’ self-health management and nurses’ work for these patients when facing future pandemics.

**Methods:** It conducted a comprehensive literature search and analysed relevant texts. This article reviews the daily health management of diabetic patients in the context of the COVID-19 pandemic and the role that nurses should play in this process. We used Pubmed to search for literature related to the role of nurses and self-management of health during COVID-19, which were subsequently used as seed literature for a complementary snowball method search.

**Results:** Nine publications were retrieved from the PubMed database with a cumulative total of 5833 adults. An additional 28 additional literature were retrieved using the snowball method using these seed literature. Diabetic patients face challenges in self-health management during the pandemic, including disruptions in healthcare access, increased mental problems, and unhealthy lifestyles. Nurses serve as the front-line interface between these patients and the healthcare system. Adopting telehealth and remote consultation has effectively bridged the gap created by social distancing measures.

**Conclusion:** Daily self-health management can significantly improve glycemic control and reduce the risk of diabetes-related complications, which is vital during the pandemic when patients may be experiencing disruptions to their routine care. Moreover, empowering patients through educational initiatives led by nurses can bring better self-monitoring, medication adherence, and lifestyle modifications, all of which are crucial in mitigating the effects of diabetes on the body’s immune response, thus reducing the severity of COVID-19 if contracted.

## 1. Introduction

Diabetes has become a growing health problem worldwide, with the number of diabetic patients increasing significantly over the past 35 years (1). According to the International Diabetes Federation (IDF), approximately 425 million adults (20-79 years old) had diabetes in 2017, and this number will increase to 629 million by 2045 (1). The report also shows that diabetes prevalence has risen faster in low- and middle-income countries than in high-income countries over the past decade (1). Adults diagnosed with diabetes are 3.5 times more likely to be hospitalised than adults without a history of diabetes, while those with prediabetes are 1.3 times more likely to be hospitalised (1).

However, the COVID-19 pandemic has presented unique challenges for individuals with diabetes, especially for patients with diabetes comorbidities, as they are at a higher risk of severe illness if infected with the virus (2). The number of diabetic patients ranks second among COVID-19 patients, second only to cardiovascular and cerebrovascular diseases (3). Therefore, people with diabetes are at a higher risk of contracting COVID-19, and infection may worsen their condition. One study reported that among COVID-19 patients, those with diabetes are twice as likely to be admitted to the intensive care unit (ICU) as those without diabetes (4). Maintaining reasonable blood sugar control will help protect people with diabetes against COVID-19 infection. However, with the global spread of COVID-19, epidemic prevention policies in various places, such as home quarantine and community blockade, have brought many inconveniences to the blood sugar control of diabetic patients, such as difficulty in seeking medical treatment, limited medical resources, and difficulty in regular diet and daily life (5).

In addition, the COVID-19 epidemic poses severe challenges to nurses in properly comforting, guiding, and diagnosing patients with diabetes to control their daily blood glucose levels (2,6). The role of nurses in supporting diabetic patients has never been more significant, as they play a vital role in educating, guiding, and advocating for these individuals. Therefore, based on the background of the COVID-19 epidemic, we discuss issues such as how to reasonably manage blood sugar control and testing, dietary intake, daily activities, drug treatment, education, and prevention of other diseases in patients with diabetes, as well as what nurses should do in this process.

## 2. Methods

This study explores the daily self-health management of diabetic patients and the role of nurses during the COVID-19 pandemic. We aimed to identify challenges, adaptations, and effective practices in nursing care under pandemic conditions. The methodology followed a narrative literature review framework, adhering to PRISMA guidelines (7), with additional snowballing strategies to supplement the search. This mixed research methodology was gradually explored and developed by Yan Bo in the course of his past research (8–16).

### 2.1 Search strategy

A systematic search was conducted in PubMed using the following keywords and MeSH terms. The search strategy consisted of four main keywords, namely nursing, self-health management, COVID-19 and diabetes (**Table 1**).

**Table 1.**
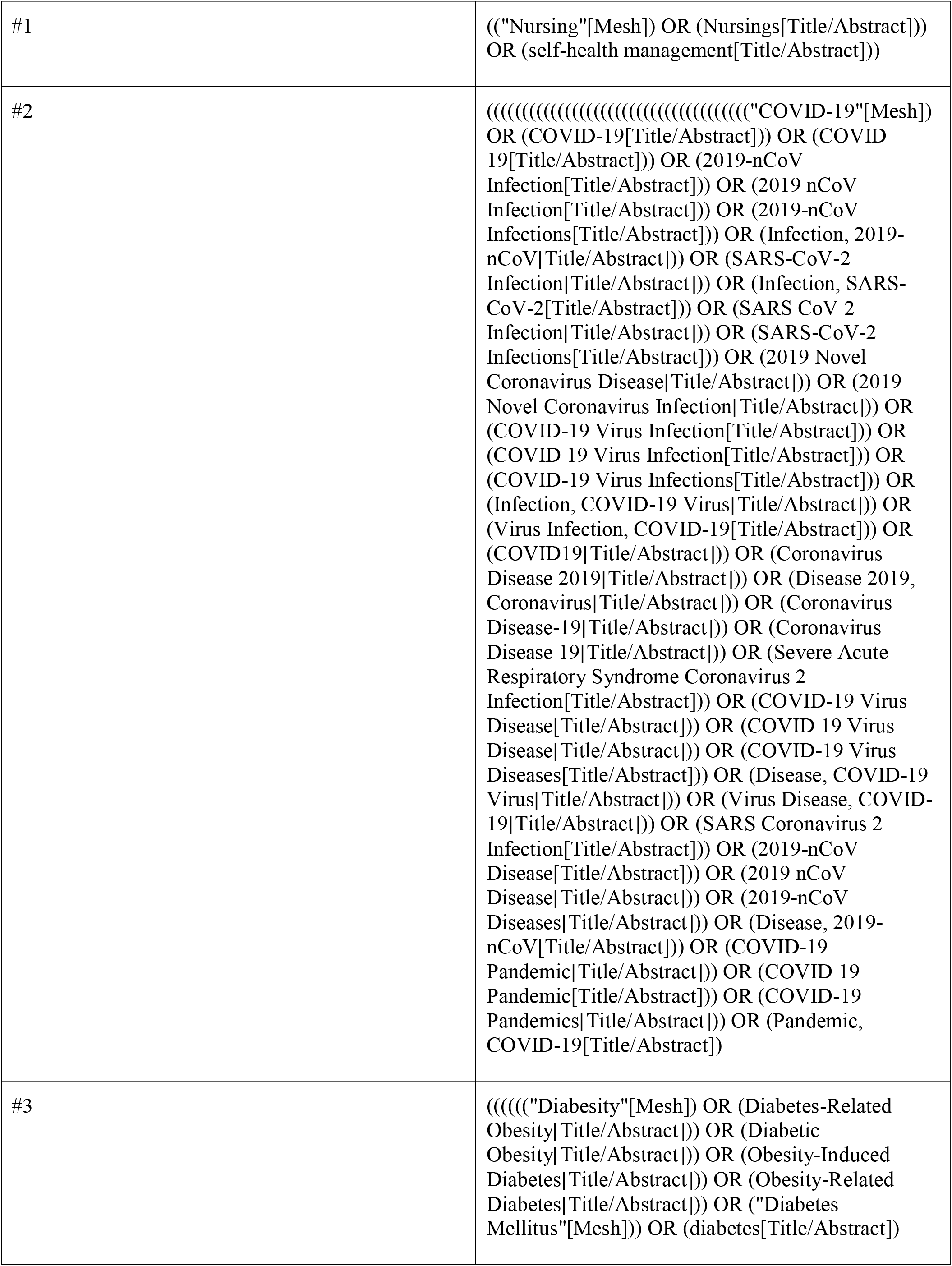
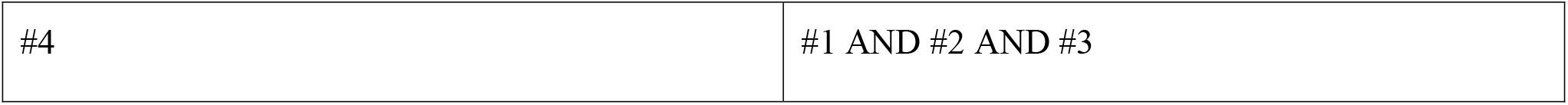
Search strategy.

All the literatures obtained from the initial search were served as “seed literatures” for the subsequent snowballing method.

### 2.2 Inclusion/exclusion criteria

Inclusion criteria: (1) Studies involving adults (≥18 years) diagnosed with diabetes (ICD-11 criteria). (2) Literature addressing self-health management practices or nursing roles during COVID-19. (3) Peer-reviewed articles published after 1 January 2020.

Exclusion criteria: (1) Studies focusing on non-diabetic populations. (2) Pre-pandemic studies or non-COVID-19 contexts.

### 2.3 Snowballing strategy

Although some studies in the literature consider it important to retrieve all the literature that needs to be included in the study at the outset (17–19), we believe that the focus on a specific topic should be on step-by-step reasoning rather than being comprehensive and systematic. Based on this consideration, we will incorporate the retrieved literature related to self-management of diabetic patients during COVID-19 in the PubMed database according to the search strategy beforehand. More relevant literature is then progressively included using a snowballing strategy based on the need for stepwise reasoning. The snowballing strategy Yan Bo was previously successfully applied in investigating the public’s health needs for an AI platform during COVID-19 (20). Based on the stepwise inference process using the snowballing method to obtain literature, which Yan Bo has used in previous studies (10,11). This strategy is more likely to find critical literature than random or comprehensive searches.

Since the snowballing method of obtaining literature was developed based on a previous study by Yan (20). Sufficient detail will be given here. We had to take the lead in developing strategies that would allow us to obtain, on the first attempt, precise literature related to the role of nurses and self-health management for people with diabetes during COVID-19. These first-time acquisitions will serve as seed literature for the snowball method. In bibliographic method studies, the method of obtaining precise literature is generally an intentional search for specific keywords, but we chose the systematic review method of retrieving literature in order to obtain a sufficiently comprehensive seed literature. The retrieved literature must be included in the study in its entirety irrespective of its size, thus ensuring a sufficient number of seed literature. This is because the literature obtained by the systematic review method is dependent on the intentionally selected keywords for a comprehensive or systematic search. Although some systematic reviews of the research reporting process have argued that only randomised controlled studies can be of interest to the researcher (17–19), for narrow topics even if a literature review presents the idea of the role of specific nurses and self-health management in people with diabetes, that would be valuable. For this reason we did not qualify the type of literature or research methods. According to PRISMA guidelines (7), a systematic review is complete once a systematic search is performed and the evidence is superimposed. The systematic review at this point, due to limitations dependent on the search strategy, only indicates systematic within the current researcher’s perspective. This may result in the inclusion of studies that are underrepresented in the literature or missing necessary details. It is therefore important to develop a methodology that allows additional access to the literature on this topic. In this study, once the literature obtained based on a systematic search has been completed with an evidence overlay, additional intentional supplementary searches can be conducted based on the different topics of the overlay evidence.

Although some studies have argued that it is important to retrieve all the literature that needs to be included in a study at the outset (17–19), we believe that the focus on a particular topic should be progressively reasoned rather than comprehensive and systematic. Based on this consideration, we will include in the PubMed database literature related to self-management in diabetic patients retrieved during COVID-19 according to a pre-established search strategy. Then, according to the need for step-by-step reasoning, a snowball strategy will be used to gradually include more relevant literature. Previously, Yan Bo had successfully applied the snowballing strategy to investigate the public’s health needs for AI platforms during COVID-19 (20). The step-by-step reasoning process based on the use of the snowballing method to obtain literature was also used by Yan Bo in his previous studies (10,11). This strategy makes it easier to find key literature compared to random or comprehensive searches.

In this study, Yan Bo describes the practical steps of the snowball method of acquiring supplementary literature. Supplementary literature was obtained through three steps after retrieving the seed literature.

#### Step 1

Backward snowballing, which means checking whether any of the references cited in the seed literature meet the previously established inclusion criteria, and supplementing them for inclusion if they do.

#### Step 2

Snowballing forward, this means discovering new articles citing the seed literature through Google Scholar and Scopus. Assuming that these new articles met the inclusion criteria, they were added.

#### Step 3

Theme word snowballing, in the process of reading the seed literature, if ideas and content were found that expanded on each other with the theme word of the study, keywords for the ideas and content were distilled and searched with intent in PubMed. For example, if a piece of literature mentions that a relevant point of view for self-health management is that people with diabetes need to self-control their diet, then self-control diet (topic word) and COVID-19 (context word) would be used as intentional search terms, and then literature obtained based on the subjective intent after searching in PubMed would be referred to as supplemental literature obtained by the snowballing method. This use of snowball method to obtain literature based on stepwise reasoning process was also used by Yan Bo in his previous studies (10,11).

The research criteria for the snowballing method of obtaining literature were aligned with the inclusion and exclusion criteria of the systematic review.

### 2.4 Data extraction

The seed literature needs to be read in full individually to extract key information: first author, study design, study population, study characteristics, references, cited literature, self-health management or care for people with diabetes.

### 2.5 Quality assessment

This study used a narrative approach to synthesise evidence, so quality of evidence assessments and publication bias assessments were not applicable. This narrative review was evaluated using the Scale for the Assessment of Narrative Review Articles (SANRA)(21).

## 3. Results

### 3.1 Literature search and study characteristics

The findings of this study highlight the importance of daily self-health management practices and nurses’ roles for diabetic patients during the COVID-19 pandemic.

A total of 9 studies were retrieved in PubMed based on a pre-established search strategy (22–30). These nine studies constituted the seed literature for the snowballing strategy. The process of narrative inference of content based on seed literature was progressively included in the flowchart for each retrieved piece of literature that required new literature (**Figure 1**). We summarised the results of all seed literature in the narrative reasoning process in a table (**Table 2**). There were three narrative reviews, two cross-sectional observational studies, three cohort studies, and one bibliometric study, totalling a cumulative total of 5,833 adult effects in these nine pieces of literature on inductive reasoning. We then superimposed the evidence on the role of nurses and self-health management using a narrative approach after categorising them according to topic, in which a rolling ball approach is used for specific topics to supplement the retrieved literature. After applying inclusion/exclusion criteria, 37 studies (9 seed; 28 supplementary) were included for analysis.

**Table 2.**
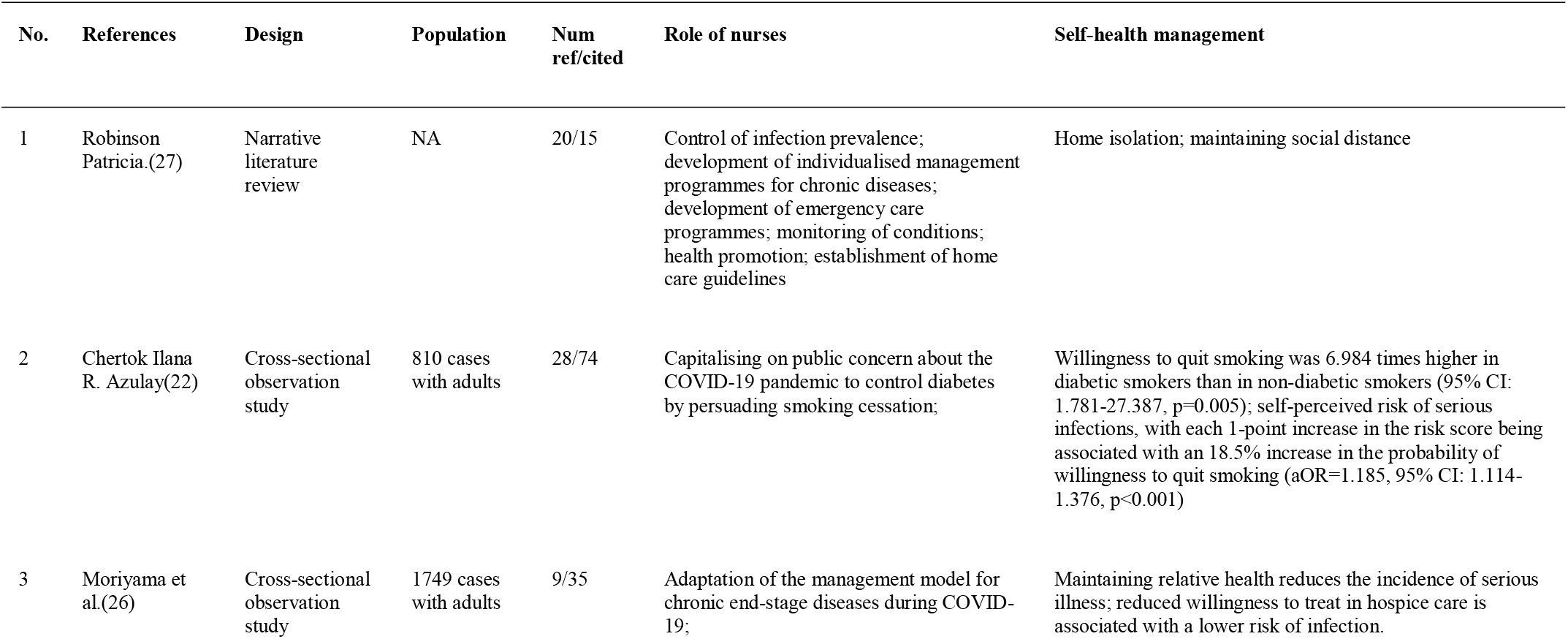
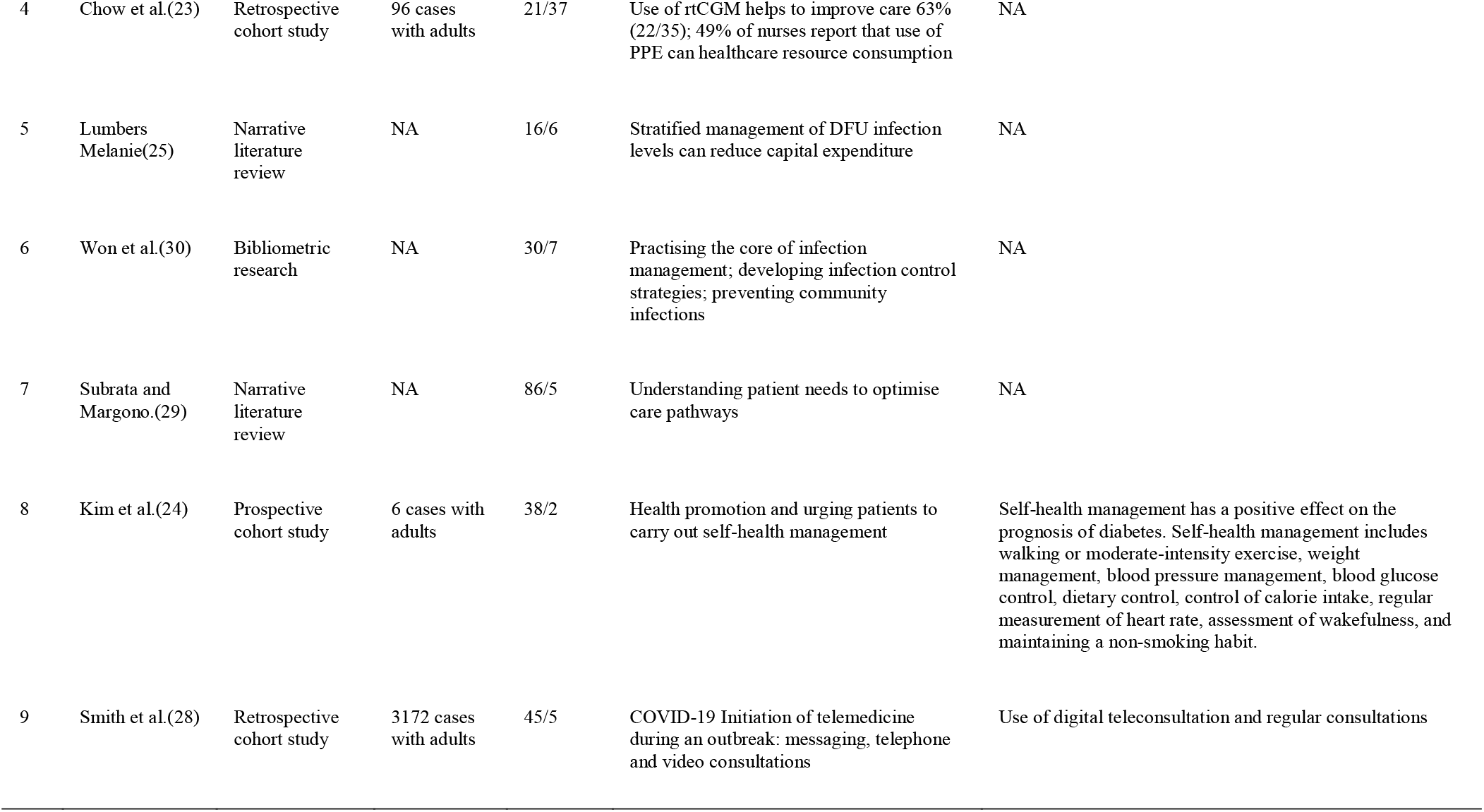
Characteristics and Key Findings of Seed Literature (n=9).

**Figure 1.**
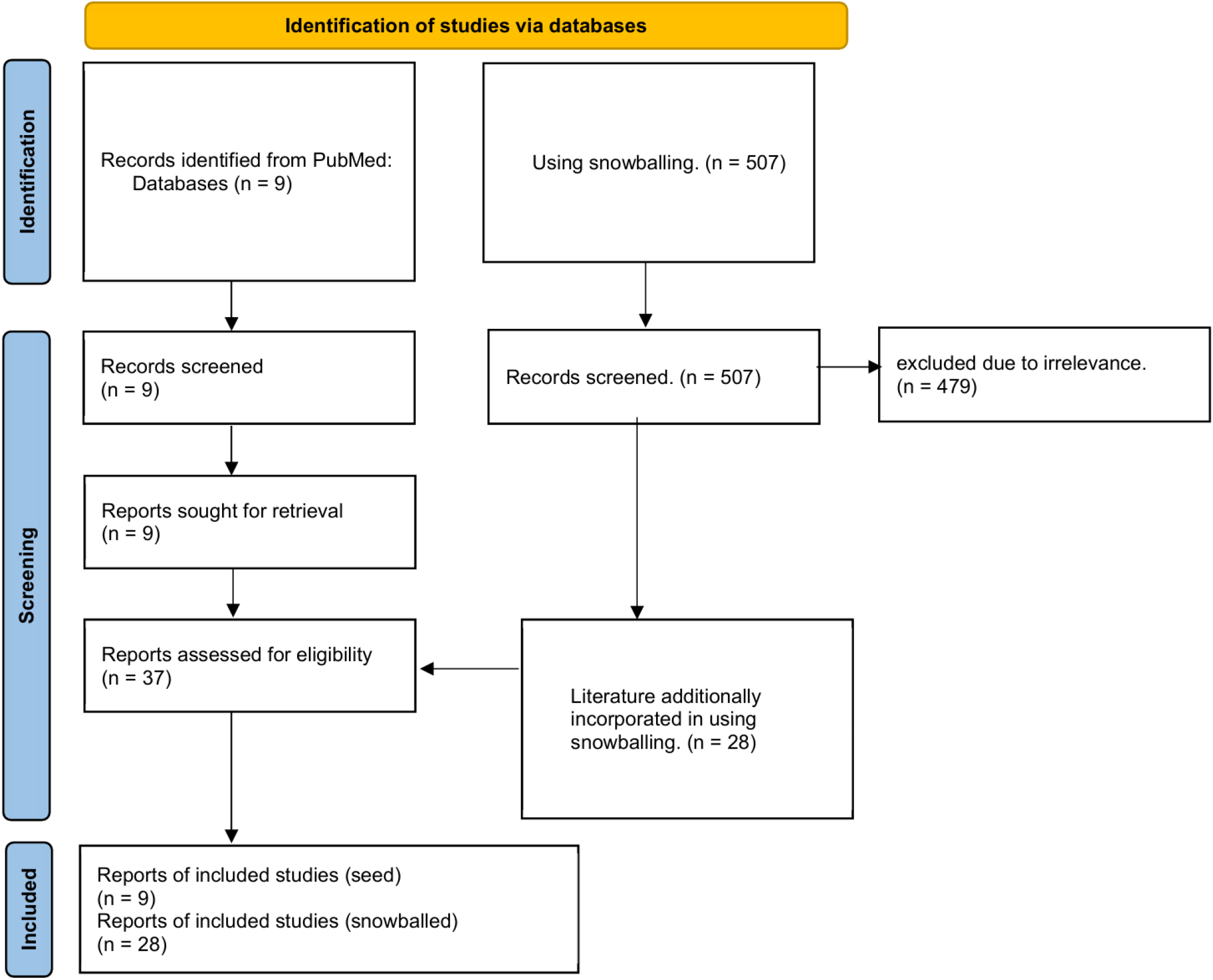
Flowchart. Identification, screening, eligibility, and inclusion stages of the review process. A total of 516 records were screened, with 37 studies meeting the final inclusion criteria.

Data were obtained from the corresponding literature (22–30). For the literature review type of studies, the role of nurses and self-health management, they were summarised using the topic distillation method. For the original research type of literature, it was summarised using correlates, risk factors or insights from the research. real-Time Continuous Glucose Monitoring (rtCGM); Personal Protective Equipment (PPE); diabetic foot ulcer (DFU); NA indicates not available.

### 3.2 Self-Health management of diabetes

According to the American Diabetes Association’s standards of diabetes care for patients and healthcare providers, diabetes care should include a comprehensive medical evaluation of comorbidities, lifestyle management, glycaemic control, pharmacotherapy, weight control, risk reduction, and prevention of diabetes complications (6). Diabetes management will be discussed considering the following standards of care:lycaemic control and monitoring, dietary intake, physical activity, pharmacotherapy, education, and prevention of COVID-19 infection in the context of the COVID-19 epidemic and some population-specific, situation-specific Diabetes Care.

#### 3.2.1 Control and detection of blood sugar

During the COVID-19 epidemic, diabetic patients must maintain regular and regular blood sugar control and monitoring. However, it may be difficult for diabetic patients to monitor and control blood sugar promptly and effectively during lockdown or isolation. The main reason is the difficulty in obtaining materials, including glucose, blood glucose meters and needles (31). Therefore, diabetic patients should be advised to purchase blood glucose monitoring materials and drugs online as early as possible to take preventive measures. In addition, blood glucose monitoring for patients with diabetes can be adjusted at any time based on age, comorbidities, clinical manifestations, and other risk factors (4,31). During COVID-19, patients with diabetes can use capillary blood tests for blood glucose monitoring (32). All hospitalised COVID-19 patients with diabetes should undergo closed blood sugar control and routine blood sugar monitoring to prevent worse situations. A previous study showed that patients with COVID-19 and diabetes were in poor health after discharge (32). Therefore, blood glucose monitoring should be performed during the 4-week follow-up after discharge to avoid exposure to infection (32).

Additionally, the timing of blood glucose control and monitoring can be tailored to specific circumstances. During COVID-19, for diabetic patients who are taking oral antidiabetic drugs and have relatively stable blood sugar (or fasting blood sugar is less than 6.9 mmol/L or glycated haemoglobin HbA1c is less than or equal to 6.5%), fasting and blood sugar tests can be performed once or twice a week for postprandial blood glucose monitoring (33).

Meanwhile, for diabetic patients with poor insulin injection effect or intermittent hypoglycaemia, blood sugar should be checked at least four times a day, that is, on an empty stomach, before lunch, before dinner, and before going to bed (34). Capillary blood glucose testing should be performed immediately at the first sign or symptom of hypoglycaemia (34). All blood glucose tests should be recorded for at least three consecutive days and informed to medical staff through remote consultation so that they can receive timely and practical guidance from medical staff (34).

#### 3.2.3 Diet control

Lockdowns during the COVID-19 epidemic may affect diabetic patients’ eating habits. Limited access to fresh fruits and vegetables and easy consumption of canned or packaged foods caused high calories or fat (35). During consultations, nursing staff and nutritionists should encourage healthy, balanced diets for diabetic patients. The recommended calorie intake for obese and non-obese diabetic patients with a sedentary lifestyle is 20 kcal/kg and 22-25 kcal/kg of ideal body weight, respectively (35). Dietary recommendations should include low carbohydrate, low fat, optimal protein intake, and no missed meals. The main recommendations for food composition are 50-60% complex carbohydrates, 25-45% dietary fibre, no more than 30% fat, and 1g/kg/d (general patients) or 0.8g/kg/d (for patients with comorbidities, protein composition in patients with renal disease and proteinuria) (35). Fat use should be no more than three teaspoons daily, and sodium intake should be less than 5 grams daily (35). Patients can also use the diabetic sugar control method - the dinner plate method-half a plate of vegetables, 1/4 protein, and 1/4 complex carbohydrates (35). Additionally, they should try to avoid alcohol, smoking, and sugary sweets (35).

#### 3.2.3 Activity exercise

Although maintaining safe social distance, lockdown, and home isolation have affected the outdoor activities and exercise of diabetic patients, regular and quantitative physical exercise is essential for them, so diabetic patients can perform limited physical exercise indoors to maintain physical health (36). Recommended home exercises include, for example, a treadmill, stationary bike or jogging and resistance training (13). Training duration should be controlled to approximately 60 minutes per day, and the intensity and type of activity can be tailored to the patient’s specific condition (36). Recommended physical activities are divided into three types: aerobic, flexibility, and strength muscle (36). At least 30 minutes a day of moderate-intensity aerobic exercise, such as brisk walking, treadmill, stationary jogging or cycling, dancing, jumping, aerobics and gardening, is recommended (36). If some of the above exercises are difficult, patients can perform 2-3 times (10-15 minutes) of easy aerobic exercises (36). Flexibility exercises include stair climbing, daily household activities, or yoga exercises as stretching techniques for 15 minutes a day as flexibility training (36,37). Strength muscle exercises recommend 15 minutes of squats, push-ups, sit-ups, forward bends, or light weightlifting as daily muscle training exercises (37). Diabetic patients with a history of heart disease or hypoglycaemia should pay special attention to physical activities and perform appropriate exercises based on their physical fitness (38).

#### 3.2.4 Medication precautions

Medical staff must conduct remote consultations during the pandemic using effective and brief tools to assess patients’ medication compliance (34). Since diabetic patients have limited access to outpatient clinics during the pandemic, medical staff should also ensure that all follow-up diabetic patients have adequate drug reserves and issue adequate drug prescriptions (34). So far, there is no substantial evidence that antidiabetic drugs can be used to treat COVID-19 patients with diabetes, although some antidiabetic drugs have shown promising effects on blood sugar control. Angiotensin-converting enzyme (ACE) inhibitors and angiotensin receptor blockers (ARB) are theoretically helpful in treating COVID-19 patients; however, no experimental data supports this theory. In addition, to prevent hypoglycaemia, the dosage of sulfonylurea drugs and insulin may need to be adjusted according to the patient’s condition during medication. All patients with diabetes are informed of adverse drug reactions and informed that they should promptly report any adverse reactions. It was reported that 44.9% of non-ICU and 72.2% of ICU COVID-19 patients received glucocorticoid therapy as daily treatment. However, glucocorticoids are associated with hyperglycaemia and can induce more severe clinical manifestations. Therefore, glucocorticoids are not recommended to treat diabetes complicated by COVID-19 to prevent secondary patient harm. There are reports of using hydroxychloroquine to treat COVID-19 patients with diabetes. In the study, hydroxychloroquine exerted a hypoglycaemic effect by reducing HbA1c and hyperglycaemia. However, chloroquine agents have hypoglycaemic and immunomodulatory effects, so the physical health of all patients should be closely monitored (39). In animal experiments, hydroxychloroquine caused an increase in insulin serum levels by providing signals to cell receptors and post-receptor clearance. Therefore, all diabetic patients receiving hydroxychloroquine should be aware of contraindications to the drug, such as diabetic neuropathy and a history of seizures.

#### 3.2.5 Diabetes-related education

Due to the COVID-19 pandemic, regular hospital visits for people with diabetes are limited (40). Therefore, conducting online or remote consultations is recommended to keep in touch with medical staff. Patients with type 1 diabetes should consult an endocrinologist, and patients with type 2 diabetes are advised to consult an internal medicine specialist or general practitioner. Endocrinologists can provide consultation by sharing educational videos and e-books using smartphone applications (such as WeChat, Tencent Meeting, and SMS). A previous meta-analysis showed that telemedicine guidance via email, phone, and video for 3-60 months during the non-COVID-19 period showed a significant reduction in HbA1c (0.37%, p < 0.001) (40). Another clinical observation on remote coaching showed similar results, with a 0.31% reduction in HbA1c (p<0.001). A recent review of 46 telemedicine studies in patients with type 1 and type 2 diabetes showed significant reductions in HbA1c (0.12%-0.86% and 0.01-1.13%, respectively) (40). For the first consultation, it is recommended to use video telemedicine mode (40). The patient’s privacy or the person’s consent (guardian, caregiver, or patient) should be protected. Medical test results and prescriptions should be integrated into the medical record. In addition, attention should be paid to the patient’s medical history, allergies, and hypoglycaemia. When oedema or visible wounds appear on the patient’s feet, the patient can inform medical staff through videos or photos to judge the condition on time (36). Healthcare staff like nurses should always remind patients to practice general precautions such as hand washing, daily wearing of masks and social distancing during remote consultations.

#### 3.2.6 Prevention of COVID-19

As mentioned, people with diabetes are at higher risk of contracting COVID-19. In general, people with diabetes should adhere to social distancing and stay-at-home policies as primary prevention methods. Contact with confirmed or suspected COVID-19 patients should be avoided as much as possible. It is recommended that people with diabetes develop a separate diabetes management plan at home or when they are sick (41). All patients with diabetes should control blood sugar according to the requirements of medical staff to reduce the risk of COVID-19 infection (32). When COVID-19 infection is suspected, all patients with diabetes should go to the hospital for consultation with a doctor or nurse (32). Once patients have a fever and cough, difficulty breathing or pneumonia, a history of travel to epidemic areas, and a history of contact with COVID-19, they should be isolated and checked in time and consult medical staff (6). When they go to the hospital or clinic for treatment, they must wear a mask during the journey (32). All medical staff should also wear medical masks when in the same room with patients (32).

#### 3.2.7 Population-Specific, Situation-Specific Diabetes Care

Diabetes health management inevitably changed during the COVID-19 pandemic. It should be individualised based on the patient’s age, coexisting medical conditions, and socioeconomic circumstances. For example, elderly patients or those with chronic kidney disease may require different approaches than younger, healthier individuals (42). Therefore, specific situations require specific responses. The following sections discuss diabetes management in specific populations or situations.

For children or adolescents newly diagnosed with type 1 diabetes, it is recommended to adopt a face-to-face consultation mode (43). Type 1 diabetic patients and their families should go to the diabetes clinic to start insulin administration (34). Healthcare professionals should ensure that patients and families receive diabetes education focusing on insulin medication, signs/symptoms and management of hypoglycaemia and ketoacidosis (34). For the follow-up of patients with type 1 diabetes, ketoacidosis testing should be recommended when hyperglycaemia occurs (44).

Gestational diabetic (GDM) patients should have a face-to-face insulin consultation at their first visit (45). Patients should receive targeted education about their diabetes and current condition for proper lifestyle management (34,46). Using telemedicine to folloup with GDM patients in case minor adjustments to insulin dosage may be necessary (40).

Additionally, older patients with diabetes are more likely to experience worsening glycemic control due to elevated blood glucose (31,44). Due to limited access to care during lockdown and isolation, hyperglycaemia or hypoglycaemia may occur, stimulating blood sugar instability (31). This situation causes further aggravation of the condition of elderly patients with diabetes and the occurrence of complications, such as ketoacidosis, infection, hyperosmolar nonketotic diabetic coma, and cardiovascular and cerebrovascular diseases, which is evident in older patients with diabetes who live alone (44). It is therefore recommended that such patients always stay in touch with medical staff and seek help quickly when needed.

When a diabetic patient develops symptoms of lethargy, vomiting, chest pain, shortness of breath, or limb weakness, it should be considered an emergency (40). They should contact medical staff for help promptly (40). Additionally, patients with diabetes who have foot lesions, gangrene, severe hypoglycaemia, gastroenteritis, and any other COVID-19-related infection should be treated on a unique basis (40). In the above situations, patients should go to the hospital/clinic for treatment or admission in time. Healthcare workers should keep abreast of patients’ signs/symptoms and take initial steps to make hospital/clinical appointments (40). For confirmed COVID-19 diabetic patients receiving intensive care, blood glucose monitoring should be strengthened, and adverse drug reactions should be detected early. It has been reported that COVID-19 patients with diabetes are twice as likely to be admitted to the ICU and receive intensive care than those without the infection (4).

### 3.3 The role of nurses on diabetic patients

#### 3.3.1 Nurses as Science Popularisers

Patient education is fundamental in diabetes management. Educating patients about diabetes, the importance of diet and exercise, medication adherence, and self-monitoring of blood glucose can empower them to manage their condition effectively.

Research shows that nurses play a significant role in educating people with diabetes to manage their disease (41). Further research shows that when nurses are involved, diabetes knowledge positively impacts patients’ conditions, demonstrating the importance of nurses’ diabetes education in improving glycemic control (47). In a recent study by Bostrom, which showed the importance of nurses in patient education, diabetes specialist nurses believed that one of their roles is to “be a teacher”, that is, how to educate patients, understand the latest status of patients, inform them about the disease, possible complications, and test results (41). Therefore, nurses have a vital role in diabetes education. Wexler also proved this through a randomised trial study of two groups of patients (48). In the study, one group received usual care, and the other received formal education from nurses and specialists. The study results showed that the average blood glucose level of hospitalised patients in the nursing and expert education group was lower than that of the standard nursing group (48). One year after discharge, HbA1c reductions were more significant in the nursing and specialist education groups (48). Similarly, Raballo found that specialised care patients had better outcomes (49).

Furthermore, these findings showed that patients who received professional care had mostly positive attitudes compared to regular care (48,49). Compared with routine care, professional care patients have more reasonable and scientific self-care knowledge (49). These studies demonstrate the critical role of nurses in diabetes education and in improving patients’ glycemic control.

#### 3.3.2 Nurses as professional caregivers

According to the definition of advanced practice nurse (APN) by the UK Regulatory Council and the Nursing and Midwifery Council (NMC) (2005), skilled nurses can play a variety of roles in the care of patients with diabetes, such as deciding and implementing treatment, regular physical examinations, and ensuring that each patient’s treatment and care is based on the best, among other things (50). One element of advanced care found in the current literature is the involvement of advanced care practitioners in managing specific medications for patients with diabetes, a role typically performed by nurses when caring for hospitalised patients (51). Additionally, many studies have mentioned the role of nurses in prescribing medications. For example, Carey and Courtenay found that over two-thirds of UK specialist nurses prescribe medication for common complications of diabetes, including hypertension, hyperlipidaemia, and cardiovascular disease, even though they spend less than 20% of their time in the week doing so (52). This result suggests that nurses spend most of their time dealing with other nursing activities and less advising and ordering medications. It also found that nurses’ contribution was limited to adjusting patients’ medications as it showed that only 17.5% of nurses reported participating in this work in 1990, while only 15.6% reported participating in 1999 (52). A recent study further supported this result, which found that 77% of UK hospitals had one or more nurses who had attended a nurse-prescribing course (53). Only 48% were involved in patient medication adjustments (53). Therefore, although nurses have had advanced knowledge and skills in medication management of patients with diabetes over the years, their effectiveness in the actual process still has limitations.

Additionally, the literature highlights the importance of nurses in screening for diabetes and its complications, which is the remit of APNs (53). Research shows that nurses primarily screen for eye and foot complications and brief physicians on complications or problems (42). Another essential role of the APN in caring for diabetic patients is that of collaborator. The primary characteristic is to play the physician assistant role, such as helping doctors provide adequate care (54,55). In other words, nurses mainly carry out nursing activities according to doctors’ orders rather than popularising relevant knowledge to patients. In addition, there is evidence that doctors often make the following decisions based on nurses’ initial assessment of patients, suggesting that nurses act as intermediaries between dotors and patients, introducing patients’ complications or problems to doctors and helping doctors diagnose and treat them (54). In addition, nurses organise and plan diabetes care among themselves, physicians, and other professionals who work together to care for diabetic patients (54).

#### 3.3.3 Nurses’ psychological counselling role for diabetic patients

The literature suggests that nurses play a motivating role for patients with diabetes during the pandemic. Some studies have shown the importance of nurses in providing psychological comfort to patients with diabetes. Rita found that nurses’ psychological comfort substantially improves diabetic patients’ self-care and control during the pandemic, and psychosocial specialists often refer patients to nurses (56). Furthermore, although nurses provide excellent care, it still needs to be improved compared to patients’ care needs (56). Another study further supports this result and found four common strategies nurses use to encourage patients: education to enhance self-care, advocacy and reflection on self-care actions, valuing social relationships, and humanising complexity (56).

## 4. Discussion

One of the critical discussions surrounding daily self-health management is the importance of glycemic control. Diabetic patients must monitor their blood glucose levels regularly and adhere to their medication regimens to maintain stable glycemic control. Although medication adherence and blood glucose monitoring were reported to be high, there is room for improvement in engaging in regular physical exercise, following a healthy diet, and practising self-management activities. However, the pandemic has disrupted routine healthcare visits, making it challenging for patients to access regular check-ups and monitoring. Nurses have stepped in to bridge this gap by providing virtual or telehealth consultations, allowing for remote monitoring and support. This approach has proven effective in ensuring continuity of care and minimising the risk of exposure to the virus for both patients and healthcare professionals.

Another essential aspect of daily self-health management is the promotion of healthy lifestyle practices, including regular physical activity, healthy eating habits, and stress management techniques. Nurses have actively provided resources and guidance to diabetic patients, helping them adopt these behaviours and make sustainable lifestyle changes. By emphasising the importance of self-care, nurses empower patients to take control of their health and reduce the risk of complications associated with diabetes.

Furthermore, the role of nurses as advocates for diabetic patients cannot be overstated. Their role in supporting diabetic patients during this challenging time was found to be crucial, emphasising the need for ongoing education, counselling, and monitoring. They have been instrumental in coordinating care with other healthcare providers, ensuring patients access to essential medications and supplies. Nurses have also proactively addressed any barriers or challenges patients face in managing their condition. This phenomenon includes addressing financial constraints, transportation issues, or difficulties accessing healthcare facilities due to lockdowns or restrictions. By advocating for their patients, nurses are crucial in improving healthcare outcomes and ensuring that diabetic patients receive the necessary support and resources to manage their condition effectively.

However, acknowledging that the pandemic has significantly strained healthcare systems, including nursing staff, is essential. Nurses have had to adapt to new protocols, navigate increased workloads, and manage their well-being amidst the pandemic. It is crucial to provide support and resources to nurses to ensure they can continue providing optimal care to diabetic patients.

These findings have implications for nursing practice, suggesting the need for targeted interventions to promote self-health management practices and enhance the role of nurses in diabetes care during present and future pandemics.

### Implications of the study

Help develop strategies for self-health management of diabetes; confirm the role of nurses in the above process.

### The added value of research

Incorporating robust daily self-health management routines, coupled with the dedicated support of nurses, can be profound in maintaining and improving the health outcomes of diabetic patients during the COVID-19 pandemic. As the healthcare landscape evolves, these insights can help inform best practices and shape interventions to safeguard patients’ well-being with chronic conditions amidst future public health threats.

## 5. Conclusions

Therefore, based on the above review, the health management of patients with diabetes should receive corresponding attention during the COVID-19 pandemic, which reminds patients and their families of issues they should notice in daily health management. These methods can be a reference in possible future infectious disease pandemics and not just for COVID-19. We should also focus on the special conditions of diabetic patients and formulate unique care plans to respond to the changing external environment and disease development at any time.

In conclusion, daily self-health management is crucial for diabetic patients, especially during the pandemic. Nurses are essential in supporting these individuals by providing education, virtual consultations, promoting self-care practices, and advocating for their needs. The evidence highlights the profound impact of nurses’ role in diabetic patients during the pandemic. It emphasises the need for comprehensive education and the implementation of telehealth services to ensure high-quality care when encountering unprecedented challenges. Their expertise and dedication are invaluable in ensuring diabetic patients receive the necessary care and support to manage their condition effectively during these unprecedented times.

## Supporting information

SANRA

## Data Availability

All data produced in the present work are contained in the manuscript

