## Supplementary material for "Self-health Management and Role of Nurses of Diabetic Patients: A systematic review during COVID-19 Pandemic": SANRA

### Scale for the Assessment of Narrative Review Articles – SANRA

Please rate the quality of the narrative review article in question, using categories 0–2 on the following scale. For each aspect of quality, please choose the option which best fits your evaluation, using categories 0 and 2 freely to imply general low and high quality. These are not intended to imply the worst or best imaginable quality.

#### 1) Justification of the article's importance for the readership

| Criterion | Score | Rating |
|---|---|---|
| The importance is not justified. | 0 | 2 |
| The importance is alluded to, but not explicitly justified. | 1 | |
| The importance is explicitly justified. | 2 | |

#### 2) Statement of concrete aims or formulation of questions

| Criterion | Score | Rating |
|---|---|---|
| No aims or questions are formulated. | 0 | 2 |
| Aims are formulated generally but not concretely or in terms of clear questions. | 1 | |
| One or more concrete aims or questions are formulated. | 2 | |

#### 3) Description of the literature search

| Criterion | Score | Rating |
|---|---|---|
| The search strategy is not presented. | 0 | 1 |
| The literature search is described briefly. | 1 | |
| The literature search is described in detail, including search terms and inclusion criteria. | 2 | |

#### 4) Referencing

| Criterion | Score | Rating |
|---|---|---|
| Key statements are not supported by references. | 0 | 2 |
| The referencing of key statements is inconsistent. | 1 | |
| Key statements are supported by references. | 2 | |

#### 5) Scientific reasoning

*(e.g., incorporation of appropriate evidence, such as RCTs in clinical medicine)*

| Criterion | Score | Rating |
|---|---|---|
| The article's point is not based on appropriate arguments. | 0 | 2 |
| Appropriate evidence is introduced selectively. | 1 | |
| Appropriate evidence is generally present. | 2 | |

#### 6) Appropriate presentation of data

*(e.g., absolute vs relative risk; effect sizes without confidence intervals)*

| Criterion | Score | Rating |
|---|---|---|
| Data are presented inadequately. | 0 | 2 |
| Data are often not presented in the most appropriate way. | 1 | |
| Relevant outcome data are generally presented appropriately. | 2 | |

---

**Sumscore** 11

### SANRA – explanations and instructions

This scale is intended to help editors assess the quality of a narrative review article based on formal criteria accessible to the reader. It cannot cover other elements of editorial decision making such as degree of originality, topicality, conflicts of interest or the plausibility, correctness or completeness of the content itself. SANRA is an instrument for editors, authors, and reviewers evaluating individual manuscripts. It may also help editors to document average manuscript quality within their journal and researchers to document the manuscript quality, for example in peer review research. Using only three scoring options, 0, 1 and 2, SANRA is intended to provide a swift and pragmatic sum score for quality, for everyday use with real manuscripts, in a field where established quality standards have previously been lacking. It is not designed as an exact measurement of the quality of all theoretically possible manuscripts. For this reason, the extreme values (0 and 2) should be used relatively freely and not reserved only for perfect or hopeless articles.

We recommend that users test-rate a few manuscripts to familiarize themselves with the scale, before using it on the intended group of manuscripts. Ratings should assess the totality of a manuscript, including the abstract. The following comments clarify how each question is designed to be used.

#### Item 1 – Justification of the article's importance for the readership

Justification of importance for the readership must be seen in the context of each journal's readership.
Consider how well the manuscript outlines the clinical problem and highlights unanswered questions or evidence gaps – thoroughly (2), superficially (1), or not at all (0).

#### Item 2 – Statement of concrete/specific aims or formulation of questions

A good paper will propose one or more specific aims or questions which will be dealt with or topics which will be reviewed.
Please rate whether this has been done thoroughly and clearly (2), vaguely or unclearly (1), or not at all (0).

#### Item 3 – Description of the literature search

A convincing narrative review will be transparent about the sources of information on which the text is based. Please rate the degree to which you think this has been achieved. To achieve a rating of 2, it is not necessary to describe the literature search in as much detail as for a systematic review (searching multiple databases, including exact descriptions of search history, flowcharts, etc.), but it is necessary to specify search terms, and the types of literature included. A manuscript which only refers briefly to its literature search would score 1, while one not mentioning its methods would score 0.

#### Item 4 – Referencing

No manuscript references all statements. However, those that are essential for the arguments of the manuscript – "key statements" – should be backed by references in all or almost all cases. Exceptions could reasonably be made for rating purposes where a key statement has uncontroversial face-validity, such as "Diabetes is among the commonest causes of chronic morbidity worldwide."
Please rate the completeness of referencing: for most or all relevant key statements (2), inconsistently (1), sporadically (0).

#### Item 5 – Scientific reasoning

The item describes the quality of the scientific point made. A convincing narrative review presents evidence for key arguments.
It should mention study design (randomized controlled trial, qualitative study, etc), and where available, levels of evidence.
Please rate whether you feel this has been done thoroughly (2), superficially (1), or hardly at all (0). Unlike item 6, which is concerned with the selection and presentation of concrete outcome data, this item relates to the use of evidence and of types of evidence in the manuscript's arguments.

#### Item 6 – Appropriate presentation of data:

This item describes the correct presentation of data central to the article's argument. Which data are considered relevant varies from field to field. In some areas relevant data would be absolute rather than relative risks or clinical versus surrogate or intermediate endpoints. These outcomes must be presented correctly. For example, it is appropriate that effect sizes are accompanied by confidence intervals. Please rate how far the paper achieves this – thoroughgoingly (2), partially (1), or hardly at all (0). Unlike item 5, which relates to the use of evidence and of types of evidence in the manuscript's arguments, this item is concerned with the selection and presentation of concrete outcome data.
